# The clinical and economic impact of a community-based, hybrid model of in-person and virtual care in a Canadian rural setting: A cross-sectional population-based comparative study

**DOI:** 10.1101/2022.11.29.22282725

**Authors:** Jonathan Fitzsimon, Christopher Belanger, Richard H. Glazier, Michael E. Green, Cayden Peixoto, Roshanak Mahdavi, Lesley Plumptre, Lise M. Bjerre

**Author notes:** Correspondence to: Jonathan Fitzsimon. **Competing interests:** Jonathan Fitzsimon is the Medical Lead for the Renfrew County Virtual Triage and Assessment Centre (VTAC).

## Abstract

**Objectives:** To determine the clinical and economic impact of a community-based, hybrid model of in-person and virtual care by comparing health-system performance of the rural jurisdiction where this model was implemented with neighbouring jurisdictions without such a model and the broader regional health system.

**Design:** A cross-sectional comparative study.

**Setting:** Ontario, Canada, with a focus on three largely rural public health units from April 1, 2018, until March 31, 2021.

**Participants:** All residents of Ontario, Canada under the age of 105 eligible for the Ontario Health Insurance Plan (OHIP) during the study period.

**Interventions:** An innovative, community-based, hybrid model of in-person and virtual care, the Virtual Triage and Assessment Centre (VTAC), was implemented in Renfrew County, Ontario on March 27, 2020.

**Main outcome measures:** Primary outcome was change in emergency department (ED) visits anywhere in Ontario, secondary outcomes included changes in hospitalizations and health-system costs, using percent changes in mean monthly values of linked health-system administrative data for two years pre-implementation and one year post-implementation.

**Results:** Renfrew County saw larger declines in ED visits (−34.4%, 95% confidence interval -41.9% to -26.0%) and hospitalizations (−11.1%, 95% confidence interval -19.7% to -1.5%), and slower growth in health-system costs than other rural regions studied. VTAC patients’ low-acuity ED visits decreased by -32.9%, high-acuity visits increased by 8.2%, and hospitalizations increased by 30.0%.

**Conclusion:** After implementing VTAC, Renfrew County saw reduced ED visits and hospitalizations and slower health-system cost growth compared to neighbouring rural jurisdictions. VTAC patients experienced reduced unnecessary ED visits and increased appropriate care. Community-based, hybrid models of in-person and virtual care may reduce the burden on emergency and hospital services in rural, remote and underserved regions. Further study is required to evaluate potential for scale and spread.

**Trial registration:** Not applicable.

**STRENGTHS AND LIMITATIONS OF THE STUDY:**

- This study uses population-level health administrative data to investigate the empirical effects of a community-based, hybrid model of in-person and virtual care in rural, remote, and underserved communities, where access to comprehensive primary care is insufficient.
- Population-level data from administrative datasets were linked using unique encoded identifiers and analyzed at ICES, Ontario’s population health data steward.
- The intervention jurisdiction is compared with two similar adjoining jurisdictions and with the whole Province.
- Because of the relatively short time period studied — two years before the intervention and one year post — it remains to be seen whether the observed differences will persist over time.
- This study’s design does not allow firm inferences about causality; however, the observed changes are in the right temporal sequence and benefit from local comparisons of similar jurisdictions.

## INTRODUCTION

Providing robust primary care for rural, remote and underserviced communities is a serious policy challenge for governments around the world (1–3). Primary care has long been considered the foundation of a high-performing healthcare system (4,5), and attachment to a family physician offering timely access to comprehensive primary care has been associated with improved health outcomes and reductions in total healthcare costs (6). However, healthcare systems in Canada and internationally have struggled to provide their citizens with timely access to and continuity of primary care (7). Any system where there are unattached patients risks profound inequity of care across populations (8,9). Unattached residents can constitute a significant minority who either go without healthcare or resort to an urgent care clinic, walk-in centre, or emergency department (ED) for their healthcare (10,11).

Rural and remote communities face even steeper challenges to offering equitable, high levels of access to comprehensive primary care (12). Rural areas tend to be larger with more dispersed facilities, making travel distance a barrier to accessing care (13), and their smaller and more dispersed populations may not support an adequate supply of family physicians (12,14). In smaller rural communities, the retirement or relocation of a single physician can be devastating to the entire community’s health system (15–17).

In light of these challenges, care models that involve internet and telephone services, or “virtual care,” are emerging as a promising mechanism to increase access to primary care and specialist care in rural, remote, and underserviced areas (18,19). During the COVID-19 pandemic, many providers offered new virtual care options, particularly during pandemic lockdowns (20,21). Given these considerations, a growing evidence base suggests that virtual care may be effective in strengthening access to primary care in rural and remote communities (22). However, little is known of the empirical impact that care models utilizing virtual options can have on primary care (23).

To bridge this gap, this study presents an initial analysis of an innovative care model, the Virtual Triage and Assessment Centre (VTAC), during its first year of operation in the rural region of Renfrew County, Ontario, Canada. This study assesses the clinical and economic impact of VTAC on emergency services utilization and hospital admissions in Renfrew County during the first year of the COVID-19 pandemic.

## METHODS

### Study Setting and Context

Renfrew County is the largest county in Ontario, Canada, and an estimated 20% of the population have no family physician or alternative primary care provider (24). With no urgent care or walk-in clinics, unattached residents must choose between accessing care at an ED or forgoing care. When the pandemic began in early 2020, local healthcare leaders met to consider options to ensure that all residents would have access to COVID-19 testing and assessment and an alternative to an ED for health concerns that could be addressed in the community. Comparison was made to the “Out Of Hours” services available in parts of the United Kingdom, whereby patients can access care when their GP’s clinic is closed (25). These services blend telephone assessments, in-person care at specific clinic sites, and at-home care provided by mobile clinicians for vulnerable, housebound patients. The Renfrew County team saw an available synergy between family physicians and community paramedics, with close collaboration with other local healthcare services ranging from mental health to palliative care. Assessments by physicians working remotely enabled access to a much larger pool of family physicians, supported by paramedics and other allied health professionals locally for in-person assessment including COVID-19 testing. The resulting Virtual Triage and Assessment Centre (VTAC), established on March 27, 2020, promised to deliver access to care for all residents of Renfrew County, to reduce unnecessary ED attendance and 911 calls, and prevent unnecessary suffering or failure to access care when necessary (26). From March 27, 2020, until March 31, 2021, over 24,000 physician assessments were completed virtually. Community paramedics conducted over 21,000 COVID-19 tests and more than 700 home assessments to vulnerable and housebound residents, and 131 residents were added to a remote monitoring program.

### Study Design

To evaluate VTAC’s efficacy, we conducted a retrospective analysis of population-level health administrative data to examine changes in health-system utilization and costs for the catchment area of Renfrew County and District Health Unit (“Renfrew County”) and several peer jurisdictions. The data was obtained from provincial-level population health data repositories held at ICES, the Ontario data steward of these repositories. ICES is an independent, non-profit research institute whose legal status under Ontario’s health information privacy law allows it to collect and analyze health care and demographic data, without consent, for health system evaluation and improvement.

The primary comparison groups were Renfrew County’s neighbouring health units, Hastings and Prince Edward Counties District Health Unit (“Hastings and Prince Edward Counties”), and Leeds, Grenville and Lanark District Health Unit (“Leeds, Grenville, and Lanark Counties”), both rural regions with similar age, rurality, socioeconomic status, and health care service levels to Renfrew County, that did not implement a system like VTAC. We also obtained data for Ontario and the Champlain region, which includes Renfrew County, the Ottawa region, and much of eastern Ontario. Data was assessed for the time period from April 1, 2018, until March 31, 2021, covering approximately two years before VTAC began operations and one year after.

Our primary objective was to investigate changes by jurisdiction of residents’ ED visits anywhere in the province, overall and by visit subtype (e.g. method of arrival). Our secondary objectives were to investigate changes in acute hospitalizations and to examine health-system costs, both for the services listed above and for the system as a whole.

### Data Sources and Collection

We used the following administrative databases: the Registered Persons Database (RPDB), which provides basic demographic information about anyone who has ever received an Ontario health card number; the Client Agency Program Enrolment (CAPE) files, which indicate the enrolment of an individual in a program with a specific practitioner and primary enrolment model; the Discharge Abstract Database (DAD), which contains inpatient hospitalization information; the National Ambulatory Care Reporting System (NACRS), which captures information on patient visits to hospital and community-based ambulatory care, including emergency departments; the Ontario Health Insurance Plan Claims Database (OHIP), which contains most claims paid for physician services by the Ontario Health Insurance Plan; and the Postal Code Conversion File (PCCF) which converts Canadian postal codes to geographic areas and includes neighbourhood income quintiles by census metropolitan area. These datasets were linked using unique encoded identifiers and analyzed at ICES. The use of data in this project was authorized under section 45 of Ontario’s Personal Health Information Protection Act, which does not require review by a Research Ethics Board.

### Study Population

The study population included Ontarians with a birthdate on or before the index date of April 1, 2020. Individuals who died, who were greater than 105 years of age, who did not have an Ontario postal code, or were not OHIP-eligible as of April 1, 2020, were excluded (Figure 1).

**Figure 1:**
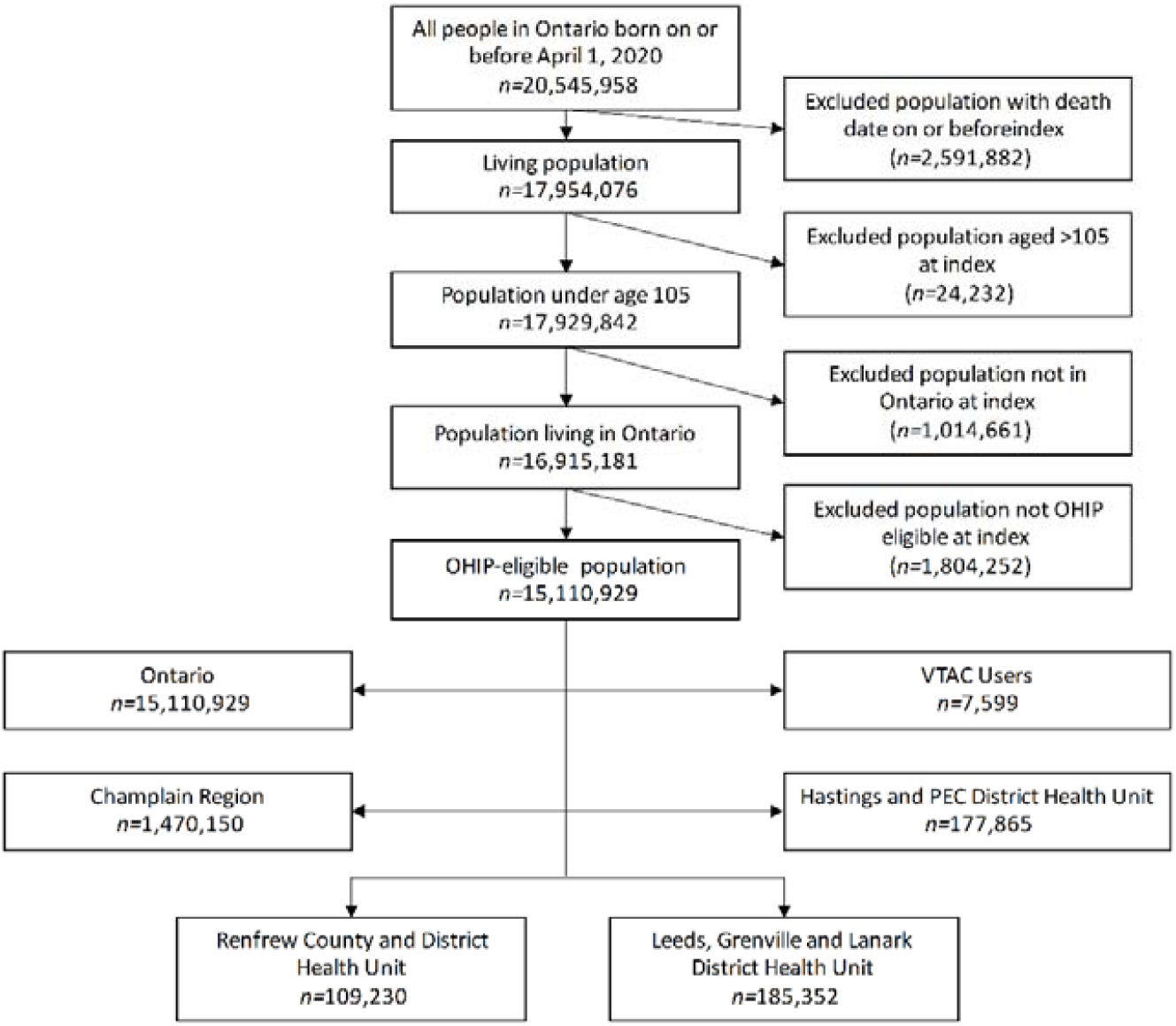
Flow diagram showing inclusion criteria for groups used in study. Note: OHIP = Ontario Health Insurance Plan.

### Patient and Public Involvement

Neither patients nor the public were directly involved in this research project.

### Statistical Analysis

Demographic information was reported for each jurisdiction including age, sex, rurality, neighbourhood income quintile, and physician rostering status. Following Statistics Canada’s recommendation, we define a community as “rural” if its population is less than or equal to 10,000 (27). Neighbourhood income quintile is based on the average neighbourhood household income, adjusted for household size and housing costs. An individual is rostered to a physician as indicated in the CAPE file. Unrostered patients may be virtually rostered or have no assigned primary care physician (see Appendix A).

We collected annual and monthly counts for our primary and secondary objectives for each jurisdiction, including unadjusted totals and rates per 100,000 residents. Percent changes in service volumes were calculated for differences between the final fiscal year, 2020/2021, and the mean of the two preceding fiscal years, 2018/2019 and 2019/2020, using SAS Enterprise Guide 7.1 (28). We calculated 95% confidence intervals for the percent changes in rates per 100,000 residents based on the ratios of mean monthly values over both time periods using version 4.0.4 of the R Language for Statistical Computing (29,30).

## RESULTS

Demographic and primary care attachment status is presented in Table 1. Renfrew County and its two neighbouring jurisdictions had generally similar population distributions in terms of age, sex, income, and primary care rostering status, with generally older, poorer, and more rural populations than the whole of the Champlain region and the province of Ontario. Although Leeds, Grenville, and Lanark Counties’ residents tended to have higher incomes and Hastings and Prince Edward Counties had a lower proportion of rural residents, Renfrew County and its two neighbouring health units were more similar to each other than to the broader Champlain region or Ontario overall.

**Table 1:** Sample population characteristics stratified by jurisdiction.

| Region; number (%) of persons |  |  |  |  |  |
| --- | --- | --- | --- | --- | --- |
| Characteristic | Renfrew County<br>and District<br>Health Unit<br>n=109,230 | Hastings & Prince<br>Edward Counties<br>Health Unit<br>n=177,865 | Leeds, Grenville<br>and Lanark<br>District Health<br>Unit<br>n=185,352 | Champlain Region<br>(LHIN 11)<br>n=1,470,150 | Ontario<br>n=15,110,929 |
| **Age, years** |  |  |  |  |  |
| 0-19 | 23,008 (21.1) | 34,054 (19.1) | 33,957 (18.3) | 311,702 (21.2) | 3,177,257 (21.0) |
| 20-39 | 24,973 (22.9) | 40,000 (22.5) | 40,271 (21.7) | 394,860 (26.9) | 4,106,872 (27.2) |
| 40-59 | 27,756 (25.4) | 46,147 (25.9) | 50,149 (27.1) | 404,665 (27.5) | 4,154,796 (27.5) |
| 60+ | 33,493 (30.7) | 57,664 (32.4) | 60,975 (32.9) | 358,923 (24.4) | 3,672,004 (24.3) |
| **Sex** |  |  |  |  |  |
| Female | 56,059 (51.3) | 90,166 (50.7) | 94,130 (50.8) | 747,641 (50.9) | 7,650,417 (50.6) |
| Male | 53,171 (48.7) | 87,699 (49.3) | 91,222 (49.2) | 722,509 (49.1) | 7,460,512 (49.4) |
| **Rurality** |  |  |  |  |  |
| Urban | 58,815 (53.8) | 113,304 (63.7) | 95,932 (51.8) | 1,306,163 (88.8) | 13,562,280 (89.8) |
| Rural | 50,415 (46.2) | 64,561 (36.3) | 89,420 (48.2) | 160,811 (10.9) | 1,510,594 (10.0) |
| Missing Data | 0 (0.0) | 0 (0.0) | 0 (0.0) | 3,176 (0.2) | 38,055 (0.3) |
| **Income Quintile** |  |  |  |  |  |
| Lowest | 32,248 (29.5) | 49,967 (28.1) | 38,347 (20.7) | 252,538 (17.2) | 2,965,949 (19.6) |
| Low | 22,008 (20.1) | 38,297 (21.5) | 33,183 (17.9) | 265,163 (18.0) | 2,957,050 (19.6) |
| Middle | 23,278 (21.3) | 33,665 (18.9) | 44,561 (24.0) | 298,376 (20.3) | 3,039,663 (20.1) |
| High | 16,964 (15.5) | 28,489 (16.0) | 44,330 (23.9) | 321,600 (21.9) | 3,053,543 (20.2) |
| Highest | 14,732 (13.5) | 27,447 (15.4) | 24,931 (13.5) | 329,256 (22.4) | 3,051,056 (20.2) |
| Missing | 0 (0.0) | 0 (0.0) | 0 (0.0) | 3,217 (0.2) | 43,668 (0.3) |
| **Primary Care Rostering Status** |  |  |  |  |  |
| Not Rostered | 18,658 (17.1) | 22,419 (12.6) | 22,786 (12.3) | 197,534 (13.4) | 1,689,219 (11.2) |
| Rostered | 74,969 (68.6) | 129,382 (72.7) | 145,134 (78.3) | 1,040,894 (70.8) | 10,884,270 (72.0) |
| Virtually | 15,603 (14.3) | 26,064 (14.7) | 17,432 (9.4) | 231,722 (15.8) | 2,537,440 (16.8) |

When we examined the group of VTAC users (n=7,599), age and income distributions were similar to those of Renfrew County, Hastings and Prince Edward Counties, and Leeds, Grenville, and Lanark Counties, but VTAC users were more likely to be female (59.4% female) and live in a rural area (57.1% rural).

### Primary Outcome: ED Visits

#### Overall ED Visits

First, we examined trends in ED visit volumes in Renfrew County compared to its neighbouring jurisdictions after VTAC was implemented. While VTAC’s implementation coincided with the widespread decrease in healthcare use during COVID-19 lockdowns (31), Renfrew County experienced a decline in ED visits that persisted to the end of the study period in March, 2021 (Figure 2).

**Figure 2:**
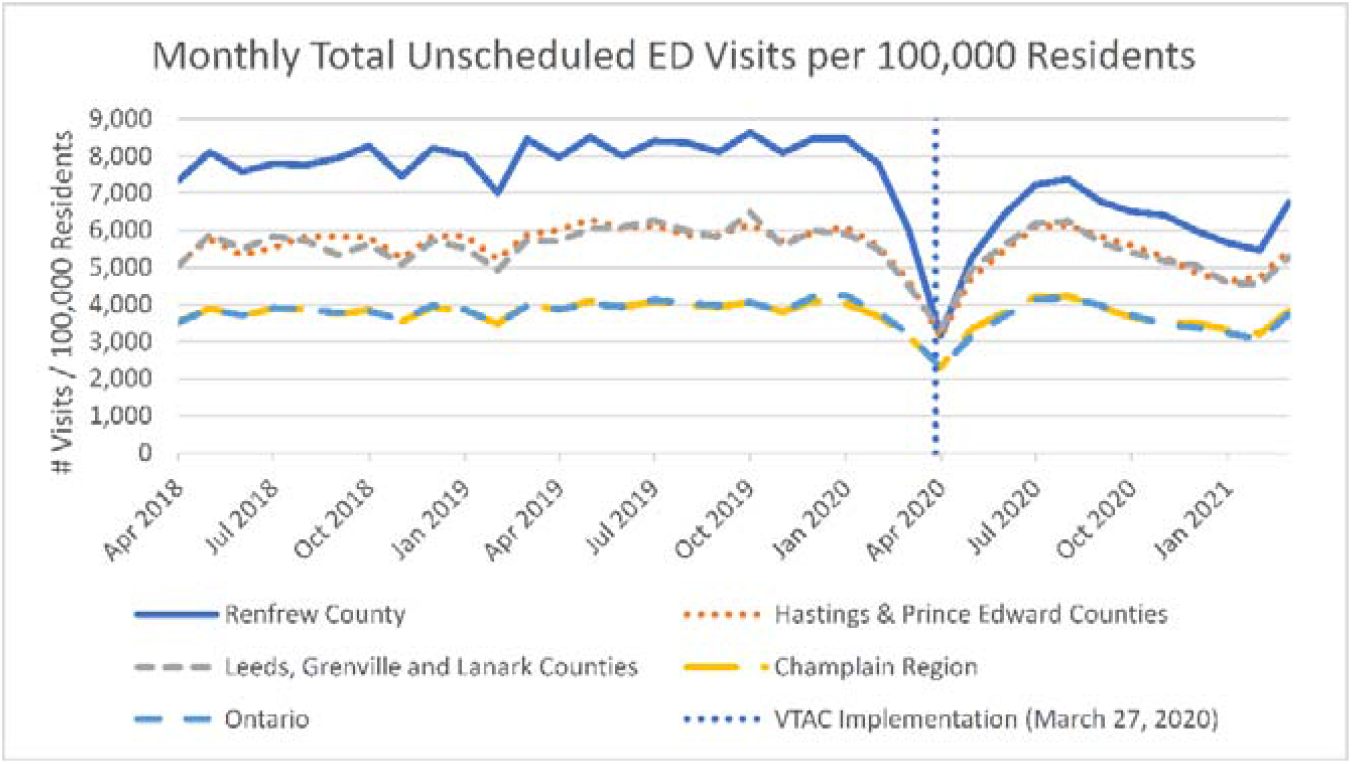
Monthly total unscheduled emergency department visits from April, 2018 until March, 2021.

ED visits in Renfrew County declined by over 34% in the year following VTAC’s implementation, more than in all other regions studied (see Table 2). On a proportional basis, this decline was more than 50% greater than those seen in any other jurisdiction studied.

**Table 2:** Unscheduled emergency department visits from 2018/19 to 2020/21.

| Region |  | FY 2018 | FY 2019 | AVG FY18&19 | FY 2020 | % Change* | 95% Conf. Int. |
| --- | --- | --- | --- | --- | --- | --- | --- |
| Renfrew County and District Health Unit | Total | 86,559 | 87,469 | 87,014 | 57,068 |  |  |
|  | Per 100k residents | 79,245 | 80,078 | 79,661 | 52,246 | -34.4% | (-41.9%, -26.0%) |
| Hastings & Prince Edward Counties Health Unit | Total | 101,754 | 103,937 | 102,846 | 80,377 |  |  |
|  | Per 100k residents | 57,209 | 58,436 | 57,822 | 45,190 | -21.8% | (-29.6%, -13.2%) |
| Leeds, Grenville and Lanark District Health Unit | Total | 105,405 | 108,735 | 107,070 | 85,096 |  |  |
|  | Per 100k residents | 56,867 | 58,664 | 57,766 | 45,910 | -20.5% | (-28.1%, -12.1%) |
| Champlain | Total | 547,869 | 551,051 | 549,460 | 446,646 |  |  |
|  | Per 100k residents | 37,266 | 37,483 | 37,374 | 30,381 | -18.7% | (-26.0%, -10.7%) |
| Ontario | Total | 5,693,888 | 5,806,200 | 5,750,044 | 4,581,053 |  |  |
|  | Per 100k residents | 37,681 | 38,424 | 38,052 | 30,316 | -20.3% | (-27.8%, -12.1%) |

\*All percentage changes are between average values for fiscal years 2018/19 and 2019/20 and fiscal year 2020/21.

#### ED Visits by CTAS

Next, we examined trends in ED visits based on their acuity as defined by the Canadian Triage and Acuity Scale (CTAS), which assigns visits a number from 1 to 5. While low-acuity CTAS 4 and 5 visits declined across all jurisdictions, the relative declines in Renfrew County were larger (Table 3). Notably, Renfrew County’s decline in non-urgent CTAS 5 visits was nearly double the provincial average (−41.5% in Renfrew County, -21.4% Ontario-wide). We also examined VTAC users’ ED visits and found that the proportion of most urgent visits, especially at CTAS 1, increased dramatically in this group. The absolute numbers, however, were very low.

**Table 3:** Unscheduled emergency department visits by CTAS level, % change from the average of the two fiscal years pre-VTAC (2018/19 and 2019/20) and one year post-VTAC inception (2020/21).

| CTAS Level | Renfrew | Hastings | Leeds | Champlain | Ontario |
| --- | --- | --- | --- | --- | --- |
| **1. Resuscitation and 2. Emergent** | -21.1% | -14.3% | -12.4% | -13.0% | -14.8% |
| **3. Urgent** | -23.2% | -16.6% | -19.6% | -11.4% | -15.5% |
| **4. Less Urgent/Semi Urgent** | -47.8% | -32.5% | -27.9% | -33.2% | -34.2% |
| **5. Non Urgent** | -41.5% | -36.7% | -5.0% | -24.4% | -21.4% |
| **0. Unknown/Missing** | -20.4% | -35.3% | -46.9% | -12.5% | -33.4% |

#### ED Admissions by Mode of Arrival

Compared to peer jurisdictions, Renfrew County’s ED admissions by ambulance increased less and ED admissions not by ambulance decreased more (Figure 3). Renfrew County’s larger drop in non-ambulance arrivals is consistent with its larger drops in CTAS 4 and 5 visits.

**Figure 3:**
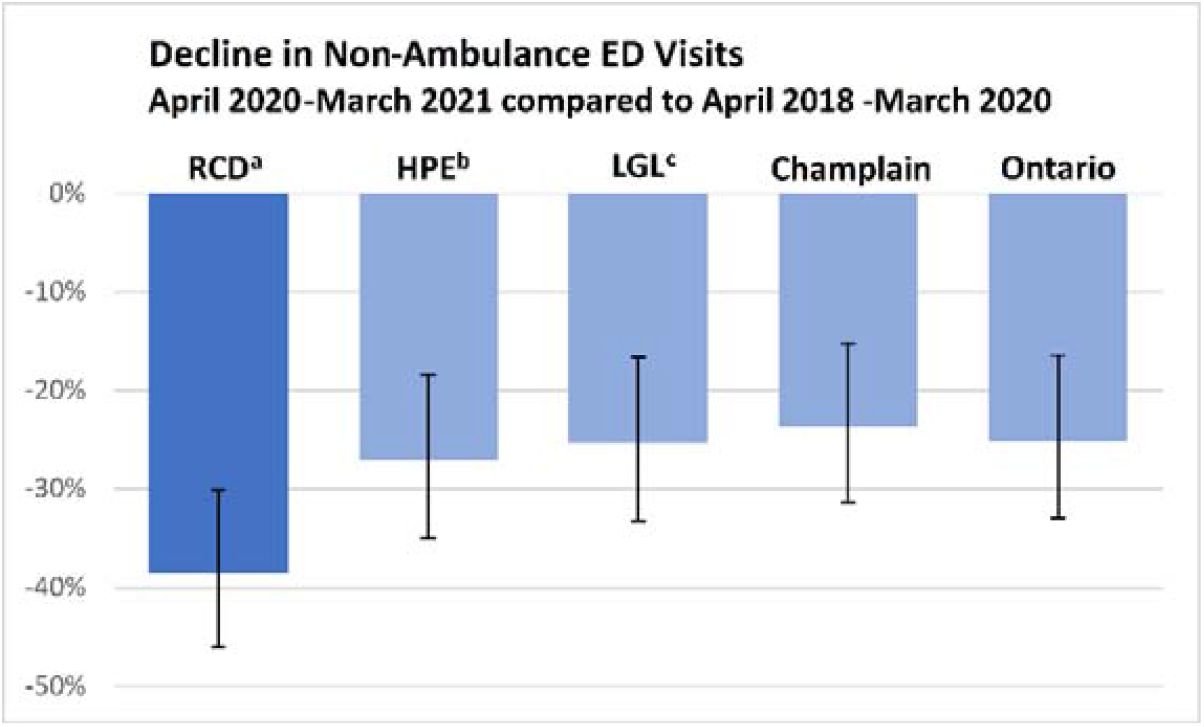
Monthly total unscheduled emergency department visits from April, 2018 until March, 2021.

### Secondary Outcome 1: Hospitalizations

The first secondary outcome we examined was hospitalizations. We found that during the study period total acute hospitalizations declined more in Renfrew County than in its neighbouring health units, and in line with larger regional trends for Champlain and Ontario (see Table 4).

**Table 4:**
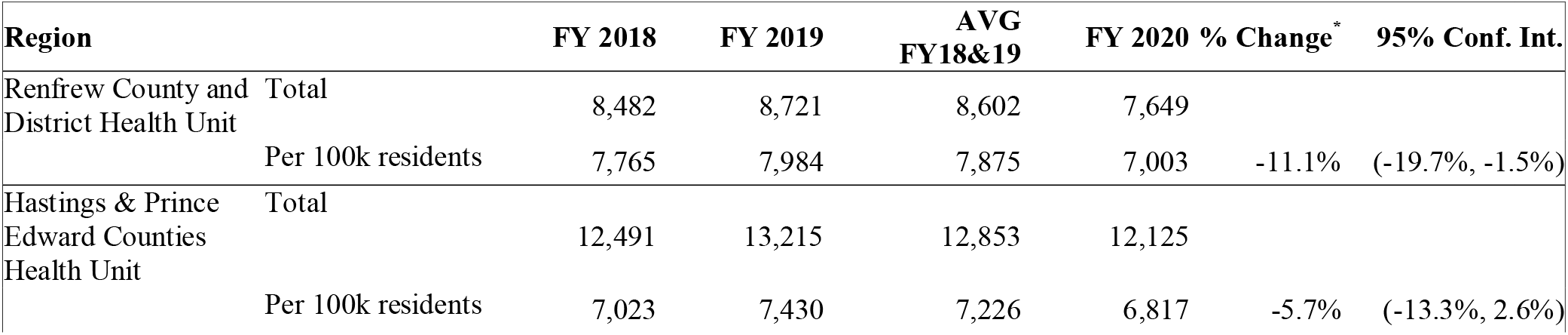
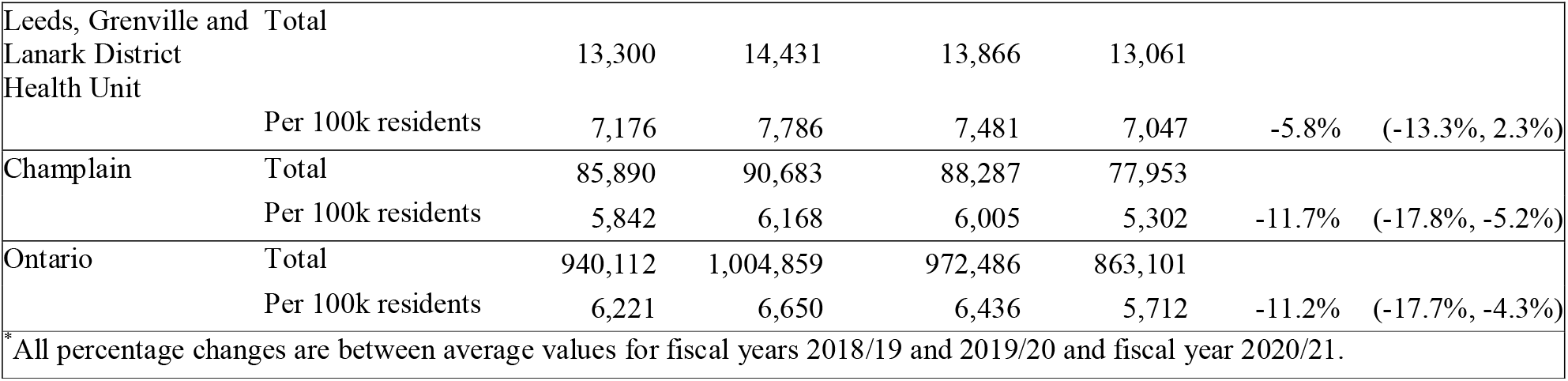
Total number of hospitalizations.

### Secondary Outcome 2: Health System Costs

Finally, we examined health-system costs, and found that costs in Renfrew County grew less than in neighbouring jurisdictions (Table 5). Renfrew County’s 2020/21 total spending per 100,000 residents was also approximately $6.5 million lower than that of its two neighbouring health units, despite all three units having near-identical average spending levels from 2018/19 to 2019/20. Renfrew County’s smaller cost increase was largely driven by lower growth in ED and hospital inpatient costs, consistent with the service volume results seen above. Note that VTAC and other regions’ COVID-19 assessment centres were funded through separate government programs and so are not reflected in these totals.

**Table 5:** Total health-system costs in Canadian dollars (costs adjusted to 2020).

| Region |  | FY 2018 | FY 2019 | AVG FY18&19 | FY 2020 | % Change* |
| --- | --- | --- | --- | --- | --- | --- |
| Renfrew County and District Health Unit | Total | 166,674,985 | 176,129,916 | 171,402,451 | 197,281,597 |  |
|  | Per 100k residents | 152,590,850 | 161,246,833 | 156,918,841 | 180,611,185 | 15.1% |
| Hastings & Prince Edward Counties Health Unit | Total | 263,238,021 | 292,843,147 | 278,040,584 | 332,919,830 |  |
|  | Per 100k residents | 147,998,775 | 164,643,492 | 156,321,133 | 187,175,571 | 19.7% |
| Leeds, Grenville and Lanark District Health Unit | Total | 280,076,601 | 301,016,063 | 290,546,332 | 346,772,601 |  |
|  | Per 100k residents | 151,105,249 | 162,402,382 | 156,753,815 | 187,088,675 | 19.4% |
| Champlain | Total | 1,839,045,450 | 1,974,784,071 | 1,906,914,761 | 2,255,531,140 |  |
|  | Per 100k residents | 125,092,368 | 134,325,346 | 129,708,857 | 153,421,837 | 18.3% |
| Ontario | Total | 20,022,014,593 | 21,606,199,097 | 20,814,106,845 | 23,958,795,043 |  |
|  | Per 100k residents | 132,500,223 | 142,983,923 | 137,742,073 | 158,552,760 | 15.1% |
| * All percentage changes are between average values for fiscal years 2018/19 and 2019/20 and fiscal year 2020/21. |  |  |  |  |  |  |

### Outcomes for VTAC Patients

VTAC users saw an 8.2% increase in ED visits at CTAS levels 1 and 2, a 13.5% increase in ED visits at CTAS level 3, and a large decline of -32.9% in low-acuity ED visits at CTAS levels 4 and 5. Acute hospitalizations increased by 30.0% for VTAC users. Details are presented in Appendix B.

## DISCUSSION

### Statement of principal findings

After implementing VTAC in March of 2020, Renfrew County’s health-system utilization changed favourably compared to neighbouring jurisdictions. Renfrew County experienced a greater reduction in ED visits, especially for less-urgent visits and non-ambulance arrival at ED, a greater decline in hospitalizations, and a slower growth in overall health-system costs when compared to Leeds, Grenville, and Lanark Counties and Hastings and Prince Edward Counties. Furthermore, VTAC users showed a large decline in low-acuity ED visits and an increase in high-acuity ED visits and hospitalizations, suggesting that unnecessary ED visits declined and medically appropriate care increased.

### Explanation of the findings

Our findings are consistent with the hypothesis that VTAC may have helped to reduce inappropriate use of emergency and hospital services and increase appropriate care in Renfrew County. Furthermore, the utilization data for VTAC users suggests a previously underserved population being diverted away from unnecessary ED visits and towards more appropriate care from a family physician. At the same time, VTAC users’ increase in urgent ED visits and hospitalizations suggests that access to timely care helped to identify emergent or urgent health issues that needed immediate treatment. Therefore, this reduction in ED visits may help to slow the future growth of Renfrew County’s health-system costs.

### Policy implications

Based on our findings, healthcare policymakers may wish to consider care models that integrate virtual options into existing local healthcare systems, including local in-person healthcare services. Our findings suggest that virtual care models can be effective at improving clinical and economic outcomes in rural, remote, and underserviced areas when they are integrated with the local healthcare system and include complementary in-person services. Due to the high degree of system connectivity in the VTAC model, the physicians delivering virtual care were knowledgeable about local resources and able to coordinate with in-person care providers, which may have enabled them to more effectively triage patients away from unnecessary ED visits and towards other appropriate care options.

### Comparison to other studies

Our findings contribute to the scientific literature in two main ways. First, we provide additional evidence that virtual care models can help to address rural health disparities (2–4), and support the emerging literature on the increased use of virtual care during the COVID-19 pandemic (5–9). Second and more importantly, we extend this literature by demonstrating the clinical and economic effectiveness of a hybrid care model that combines virtual consultations with family physicians with in-person care provided by community paramedics. Virtual care is essential to VTAC, but it is embedded in the larger regional healthcare system.

### Limitations

This study has several limitations. First, because of the relatively short time period studied — two years before VTAC and one year post — it remains to be seen whether the observed differences in Renfrew County will persist over time. The results for VTAC users should also be interpreted with caution due to its smaller sample size (n=7,599) and potential sampling bias, since VTAC users accessed healthcare by definition, whereas the other samples included all residents. In addition, other jurisdictions had COVID assessment centres, but it was not possible to do assessment centre cost comparisons. Finally, this study’s design does not allow firm inferences about causality; however, the observed changes are in the right temporal sequence and benefit from local comparisons of similar jurisdictions.

### Future directions

Future work could extend this study in several directions. First, a longer-term study of VTAC users’ health outcomes could examine the program’s downstream clinical impacts. Second, qualitative studies of patients’ and providers’ experiences with VTAC could uncover perceived barriers and enablers to achieve the program’s full potential. A qualitative study of provider feedback is underway, and a future evaluation of patient feedback is planned. Third, a longer-term analysis of VTAC’s post-pandemic effectiveness could examine its ongoing sustainability. Finally, to assess the broader transferability of our findings, programs that incorporate key elements of a community-based, hybrid model of in-person and virtual care could be implemented and evaluated in other jurisdictions.

### Conclusion

Our findings suggest that implementing a community-based, hybrid model of in-person and virtual care, was associated with a reduction in low-acuity ED visits and hospitalizations in Renfrew County, Canada, with attendant clinical benefits and cost savings. While additional research is needed to fully understand this model’s impact, our findings suggest that this model of care may be a contributor to effective health-system utilization and better health outcomes in rural, remote, and underserviced populations.

## Supporting information

STROBE Checklist

## Data Availability

No additional data available.

## Data availability statement

No additional data available.

## Ethics approval

The use of data in this project was authorized under section 45 of Ontario’s Personal Health Information Protection Act, which does not require review by a Research Ethics Board.

## Transparency statement

The lead authors affirm that the manuscript is an honest, accurate, and transparent account of the study being reported; that no important aspects of the study have been omitted; and that any discrepancies from the study as planned (and, if relevant, registered) have been explained.

## Contributorship Statement

All authors have contributed sufficiently to meet the criteria for authorship and have made the following contributions: Jonathan Fitzsimon and Lise M. Bjerre conceived of the study, and then designed the study in collaboration with Richard H. Glazier, Michael E. Green, Cayden Peixoto, and Lesley Plumptre. Roshanak Mahdavi and Christopher Belanger analyzed the data. Jonathan Fitzsimon and Christopher Belanger drafted the manuscript, and all of the authors interpreted the data, revised the manuscript critically for important intellectual content, approved the final version to be published, and agreed to be accountable for all aspects of the work. In addition, the authors take full responsibility for the data, the analyses and interpretation, and the conduct of the research.

## Acknowledgements and disclaimers

This study was supported by ICES, which is funded by an annual grant from the Ontario Ministry of Health (MOH) and the Ministry of Long-Term Care (MLTC). This study also received funding from the Ontario Ministry of Health through the INSPIRE-PHC Applied Health Research Question (AHRQ) Program. Parts of this material are based on data and information compiled and provided by: MOH, Canadian Institute for Health Information (CIHI). The analyses, conclusions, opinions and statements expressed herein are solely those of the authors and do not reflect those of the funding or data sources; no endorsement is intended or should be inferred, and views expressed do not necessarily reflect those of the Province. Parts of this material are based on data and/or information compiled and provided by CIHI. However, the analyses, conclusions, opinions and statements expressed in the material are those of the author(s), and not necessarily those of CIHI.

