## Appendix A for "The clinical and economic impact of a community-based, hybrid model of in-person and virtual care in a Canadian rural setting: A cross-sectional population-based comparative study"

### **APPENDIX A: CORE PRIMARY CARE FEE CODES**

Listed below are the fee codes for core primary care services used for virtual rostering assignment. A person is virtually rostered to a physician by retrieving core primary care fee codes in OHIP over a year 2 period (see Appendix Table A1 for fee codes). Using the standard pricing file, the physician with the highest cost is assigned to that individual as virtually rostered.

**Table A1**. Core primary care fee codes.

|  | Feecode | Description |
| --- | --- | --- |
| Primary Care Codes | A001 | MINOR ASSESS.-F.P./G.P. |
|  | A002 | Family Practice & Practice in General - Enhanced 18 month well baby visit |
|  | A003 | GEN. ASSESS. -F.P./G.P. |
|  | A007 | INTERMED.ASSESS/WELL BABY CARE-F.P./G.P./PAED. |
|  | A903 | GEN/FAM PRACT-PRE-DENTAL/OPER.ASSESS LIMIT 2 PER YEAR/PT |
|  | E075 | GERIATRIC GENERAL ASSESSMENT PREMIUM |
|  | G212 | D./T. PROC.-ALLERGY-HYPOSENSITIZATION INJECTION PLUS BASIC |
|  | G271 | D./T. PROC.-CARDIOV.-ANTICOAGULANT SUPERVISION |
|  | G372 | D./T. PROC.-INJECTIONS-INTRADERMAL/MUSCULAR ETC. EA. ADD. |
|  | G373 | D./T. PROC.-INJ. INTRADERMAL/MUSC. BASIC FEE (SHICK TEST) |
|  | G365 | D./T. PROC.-GYNAECOLOGY-PAPANICOLAOU SMEAR |
|  | G538 | D&T IMMUNIZATION-WITH VISIT, EACH INJECT. |
|  | G539 | Injection of unspecified agent - sole reason (first injection) |
|  | G590 | INFLUENZA AGENT +VISIT |
|  | G591 | Injection of influenza agent - sole reason |
|  | K005 | INDIVIDUAL CARE PER 1/2 HR |
|  | K013 | COUNSELLING-ONE OR MORE PEOPLE-PER 1/2HR. |
|  | K017 | ANNUAL HEALTH EXAM-CHILD AFT. 2ND BIRTHDAY. |
|  | P004 | OBS.-PRENATAL CARE-MINOR PRENATAL ASSESS.-SUBSEQ.PRENAT.VIS. |
|  | K130 | Periodic health visit - adolescent |
|  | K131 | Periodic health visit - adult aged 18 to 64 inclusive |
|  | K132 | Periodic health visit - adult 65 years of age and older |
|  | K030 | DIABETIC MANAGEMENT FEE |
|  | K080 | Minor assessment - Covid, Virtual |
|  | K081 | Intermediate assessment - Covid, Virtual |
|  | K082 | Primary mental health care - Covid, Virtual |
| Paediatric codes | A261 | MINOR ASSESS.-PAED. |
|  | A268 | Paediatrics - Enhanced 18 month well baby visit |
|  | K267 | ANNUAL HEALTH EXAM-CHILD-AFT. 2ND BIRTHDAY PAED. |
|  | K269 | ANNUAL HEALTH EXAM-PAEDIATRICS-ADOLESCENT-OFFICE |
