## Appendix B for "The clinical and economic impact of a community-based, hybrid model of in-person and virtual care in a Canadian rural setting: A cross-sectional population-based comparative study"

### **APPENDIX B: DATA TABLES FOR VTAC USERS**

Presented below are supplementary data tables for the group of VTAC users.

**Table B1:** Health metrics for the group of VTAC users (n=7,599).

| **Metric** | **Base** | **2018** | **2019** | **AVG FY18&19** | **2020** | **% Change** |
| --- | --- | --- | --- | --- | --- | --- |
| Unscheduled ED visits | Total | 9,507 | 10,824 | 10,166 | 9,010 |  |
|  | Per 100k residents | 125,109 | 142,440 | 133,774 | 118,568 | -11.4% |
| Hospitalizations (Acute or ALC) | Total | 806 | 939 | 873 | 1,134 |  |
|  | Per 100k residents | 10,607 | 12,357 | 11,482 | 14,923 | 30.0% |
| Total cost | Total | 14,809,008 | 16,784,302 | 15,796,655 | 25,867,440 |  |
|  | Per 100k residents | 194,881,011 | 220,875,141 | 207,878,076 | 340,405,843 | 63.8% |

**Table B2:** VTAC Users' unscheduled ED visits per 100,000 residents by CTAS level.

| **CTAS Level** | **2018** | **2019** | **AVG FY18&19** | **2020** | **% Change** |
| --- | --- | --- | --- | --- | --- |
| 1. Resuscitation and 2. Emergent | 14,620 | 15,923 | 15,272 | 16,528 | 8.2% |
| 3. Urgent | 44,545 | 52,244 | 48,395 | 54,915 | 13.5% |
| 4. Less Urgent/Semi Urgent | 45,269 | 42,321 | 43,795 | 31,267 | -28.6% |
| 5. Non Urgent | 20,490 | 31,267 | 25,878 | 15,489 | -40.1% |
| 0. Unknown/Missing | 184 | 684 | 434 | 368 | -15.2% |
